# Treating Rheumatoid Arthritis in Zanzibar: a cost effectiveness study comparing conventional, biologic, and targeted-synthetic disease modifying anti-rheumatic drugs

**DOI:** 10.1101/2023.12.05.23299489

**Authors:** Sanaa S. Said, Melf-Jakob Kühl, Bjorg-Tilde Svanes Fevang, Tone Wikene Nystad, Kjell Arne Johansson

## Abstract

To evaluate the cost effectiveness of six disease modifying anti-rheumatic drug (DMARD) treat-to-target treatment strategies for patients with rheumatoid arthritis in Zanzibar.

A Markov model was used to calculate the cost-effectiveness of various DMARD strategies in the treatment of rheumatoid arthritis over a three-year period. A health-provider perspective was used and only outpatient costs were considered. The Clinical Disease Activity Index (CDAI) was utilized for measurement of efficacy and values were obtained from literature. Quality Adjusted Life Years (QALYs) were obtained from 122 patients attending the rheumatology clinic at Mnazi Mmoja Hospital. Data on costs were obtained from the central medical stores and hospital administration. Treatment strategies were given in sequential approach based on treat to target goals of therapy. This included methotrexate monotherapy, methotrexate + sulfasalazine + hydroxychloroquine, methotrexate followed by one or two biologic/targeted-synthetic DMARDs (b/tsDMARDs). Probabilistic and one way sensitivity analysis were performed. Scenario analysis was undertaken comparing drug prices from India and Scandinavia.

Costs of therapy/patient/three years ranged from USD 634 for methotrexate monotherapy and USD 5011 for methotrexate and two consecutive b/tsDMARDs. The highest and lowest effects were 2.209 and 2.079 QALYs gained from methotrexate therapy + two consecutive b/tsDMARDs and methotrexate monotherapy, respectively. From a healthcare perspective methotrexate monotherapy was the cost-effective option at a willingness to pay of USD 282. Pairwise comparison also favored methotrexate monotherapy as the feasible option. We found that increasing the willingness to pay led to a change in the most acceptable option from methotrexate monotherapy to methotrexate followed by b/tsDMARD.

Methotrexate monotherapy is the cost-effective option for the management of rheumatoid arthritis in Zanzibar. Other options may be feasible if the willingness to pay threshold is increased or the drug prices are lowered, particularly for the b/tsDMARDs.

## Introduction

Rheumatoid arthritis (RA) is a chronic inflammatory joint disease characterized by pain, swelling, and stiffness that leads to joint destruction. In sub-Saharan African (SSA) populations, the prevalence of RA is estimated to be between 0.6 and 1.0% (1,2). With low treatment coverage, RA is included among the significantly neglected chronic disease (3). People with RA experience reduced physical functioning, quality of life, and life expectancy (4). The 2019 Global Burden of Disease estimated that RA caused 3.3 million disability adjusted life years (DALYs) and 44,000 deaths globally (5), with increasing incidence over the last decade (6).

Effective therapies for RA are available. When initiated early and aggressively, based on the treat-to-target (T2T) strategy, they improve overall outcomes and prevent disability (7-9). Recommended drugs include the affordable conventional synthetic disease modifying anti-rheumatic drugs (csDMARDS) such as methotrexate, sulfasalazine, and hydroxychloroquine, and the costlier and less accessible biological disease modifying anti-rheumatic drugs (bDMARDS) such as tumor necrosis factor alfa inhibitors (TNFi) and rituximab. Additionally, targeted synthetic DMARDs (tsDMARDS), e.g., the JAK-inhibitors tofacitinib and baricitinib, have recently become widely available (10). These therapies are available in high income countries but are limited in low- and low-middle income countries (LLMICs) (11).

While economic evaluations of advanced RA treatments for high-income settings are available (12-14), only few exist for LLMICs (15,16). Recent cost-effectiveness analyses from high-income countries focused on costly bDMARDs (17,18), which may face implementation challenges in resource constrained settings compared to relatively basic treatments such as methotrexate and triple therapy. In such settings, the opportunity costs are extremely high and fair priority setting of RA management alongside essential and very cost-effective services like basic obstetric care is important. Therefore, policy-relevant economic evaluations for SSA should compare feasible RA treatment options in regionally contextualized analyses.

Although one third of RA patients eventually require biologic therapy (19), the cost-effectiveness of biologic/targeted synthetic disease modifying anti-rheumatic drugs (b/tsDMARDs) in African settings is not yet researched and the drugs are often unavailable due to their relatively high treatment costs and fragile price negotiation systems (11). The Disease Control Priorities-3 (DCP3) review estimated the cost effectiveness of DMARD therapy in 2001 for developing countries based on available literature from Western populations. They reported possible effectiveness for corticosteroids at low doses, and for combination therapy of methotrexate, sulfasalazine, and prednisolone. For bDMARD therapy, the costs were considered prohibitive (20).

This study aims to evaluate the cost-effectiveness of six DMARD treat-to-target treatment strategies for patients with rheumatoid arthritis in Zanzibar. The strategies include combinations of methotrexate, sulfasalazine, hydroxychloroquine, prednisolone, and b/tsDMARDs. Some strategies may be more commonly practiced in resource limited settings (21,22) while others are practiced in developed countries (23, 24) where b/tsDMARDs are more readily available.

## Methods

### Model

Six Markov models were developed to compare the cost-effectiveness of the six RA treatment strategies using TreeAge® Pro Healthcare 2022 (Fig 1). Results were reported according to the 2022 Consolidated Health Economic Evaluation Reporting Standards (CHEERS) statement (25). We used a validated model (26) similar to that of Schipper et al (27) with an additional death state to include patients who died during the modelling period (S1 Fig).

**Fig 1:**
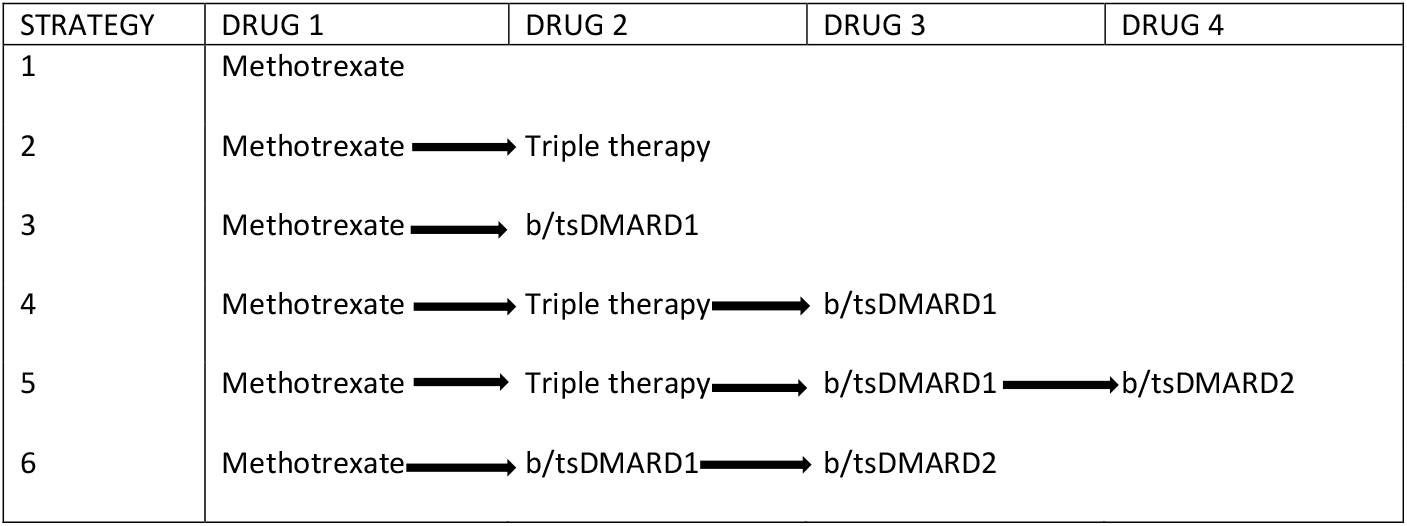
Treatment algorithm of the six included strategies. Each treatment strategy is initiated with methotrexate as the first-line drug. The arrows depict the next drug option if patients did not achieve treatment target. In all arms patients are placed on rescue therapy (methotrexate+daily prednisolone <10mg) if treatment goal was not reached. Triple therapy indicates combination of methotrexate+sulfasalazine+hydroxychloroquine, b/tsDMARD – biologic/targeted synthetic Disease Modifying Anti-Rheumatic Drug.

The Markov design was used to evaluate the differences in transitions between five health states for each strategy (Fig 2). We used Markov cycle lengths of six months, reflecting international guidelines on treatment duration before drug change (23, 28). We further defined a time horizon of three years based on the assumption that patients’ disease is less likely to show much change to available therapy after this period.

**Fig 2:**
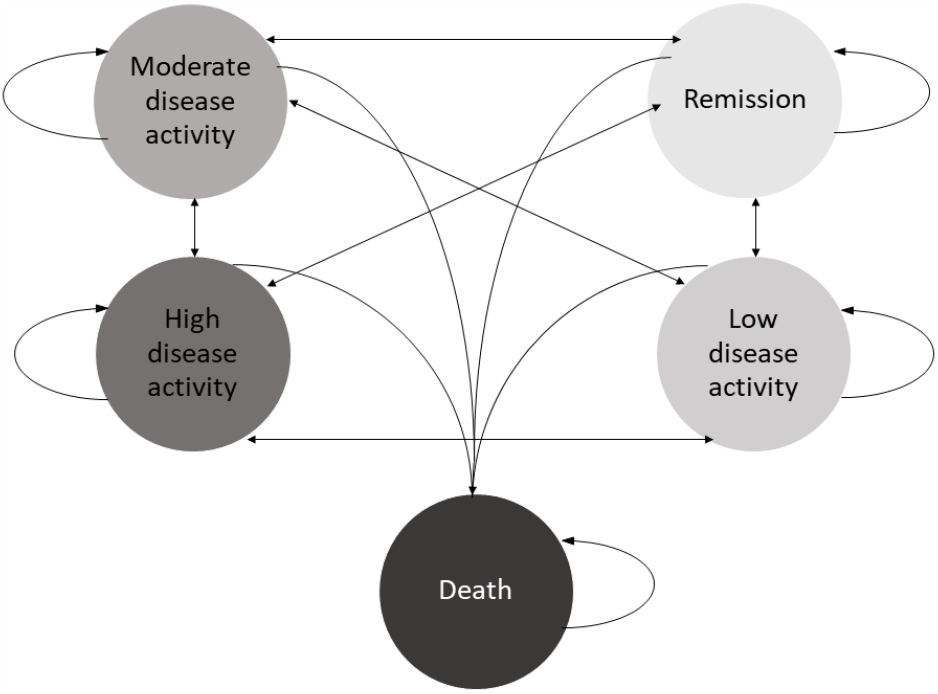
Illustration of the transition possibilities between the health states used in the model.

Based on primary data (see Study Population, below), we determined that the modelled cohort started with moderate disease activity at baseline. Disease remission or low disease activity were the desired health states (treatment target), death was an absorbing state. At the end of each cycle, disease activity was determined. If the treatment target was achieved, patients remained on the same drug therapy in the subsequent cycle. If treatment target was not achieved, they progressed to the next drug option within the same strategy. Those who failed to achieve target on all available drugs within the strategy were placed on methotrexate with low dose prednisolone (<10mg/day), termed rescue therapy.

### Interventions and costs

We determined Methotrexate as the comparator because it is the recommended first line DMARD, by both European and American guidelines unless contraindicated (24,25). Costs included in the analysis are shown in Table 2. All cost data collected were converted into USD using the exchange rates of December 2021. We assumed a healthcare provider perspective to estimate the direct and indirect costs of RA interventions, limited to outpatient clinic visits. These included four clinic visits during the first six months of therapy: at diagnosis, at one month, at three months and at six months. After this, patients who reached treatment target had one clinic visit per cycle and those not at target (moderate or high disease activity) required two visits per cycle. Costs of adverse drug events or inpatient care were not considered.

**Table 1:** Key costs for the management of RA in Zanzibar.

| Item | Zanzibar | Range |  |
| --- | --- | --- | --- |
|  | Unit costs, 6-months (USD) | Low | High |
| Methotrexate (+folic acid)<sup>1</sup> | 45 | 22.5 | 67.5 |
| Bridging therapy<sup>2</sup> | 6 | 3 | 9 |
| Sulfasalazine<sup>1</sup> | 127 | 63.5 | 190.5 |
| Hydroxychloroquine<sup>1</sup> | 111 | 55.5 | 166.5 |
| 1<sup>st</sup> b/tsDMARD<sup>3</sup> | 94* | 47 | 141 |
| 2<sup>nd</sup> b/tsDMARD<sup>3</sup> | 5456* | 2728 | 8184 |
| Prednisolone<sup>4</sup> | 20 | 10 | 30 |
| Costs at diagnosis<sup>5</sup> | 125 | 62.5 | 187.5 |
| Investigations to continue csDMARDs (1<sup>st</sup> cycle) | 57 | 27.5 | 82.5 |
| Investigations when starting b/tsDMARDs | 157 | 77.5 | 232.5 |
| Costs to continue csDMARD therapy<sup>6</sup> | 25 | 20.5 | 61.5 |
| Costs to continue b/tsDMARDs<sup>7</sup> | 56 | 3.5 | 10.5 |
| Clinic visit costs (per visit)<sup>8</sup> | 4 | 2 | 6 |

USD- United States dollar, csDMARDs – conventional synthetic disease modifying anti-rheumatic drugs, b/tsDMARD- biologic/targeted-synthetic disease modifying anti-rheumatic drugs. All costs are aggregated for 6 months except for clinic visit costs.
<sup>1</sup> Costs for hydroxychloroquine 400 mg/day, sulfasalazine 2000 mg/day and methotrexate 25 mg/week (+folic acid 5mg once weekly)
<sup>2</sup> Costs of prednisolone at 15mg tapered over 6 weeks, calcium + vitamin D supplement and omeprazole.
<sup>3</sup> Approximate prices for conventional dosage of the two cheapest b/tsDMARD in 2022 administered at recommended adult doses for rheumatoid arthritis therapy.
<sup>4</sup> Prednisolone as rescue therapy at 5mg/day. Used with Calcium + Vitamin D which contribute USD 18 to the total.
<sup>5</sup> Costs of investigations required to diagnose, start therapy, and contraception.
<sup>6</sup> If a patient is at target, this will only require investigations once/cycle but if not at target will require twice/cycle.
<sup>7</sup> Costs of pneumococcus vaccine, prophylaxis with cotrimoxazole, isoniazid preventive therapy and contraception as well as screening for pulmonary tuberculosis with chest radiograph.
<sup>8</sup> Costs of staff, building, supplies and utilities per patient per clinic visit.

**Table 2:** Transition probabilities of health states on different drug therapies for six months for patients with RA in Zanzibar.

| Intervention | Health state | Transition probability<sup>1</sup> | Range |  | Source |
| --- | --- | --- | --- | --- | --- |
|  |  |  | High | Low |  |
| Methotrexate | Remission | 0.09 | 0.0675 | 0.1125 | (32,33) |
|  | LDA | 0.34 | 0.255 | 0.425 |  |
|  | MDA | 0.38 | 0.285 | 0.475 |  |
|  | HDA | 0.19 | 0.1425 | 0.2375 |  |
| Triple therapy | Remission | 0.10 | 0.75 | 1.25 | (34) |
|  | LDA | 0.41 | 0.3075 | 0.5125 |  |
|  | MDA | 0.33 | 0.2475 | 0.4125 |  |
|  | HDA | 0.16 | 0.12 | 0.20 |  |
| b/tsDMARD 1 (early)<sup>2</sup> | Remission | 0.12 | 0.15 | 0.18 | (35) |
|  | LDA | 0.46 | 0.345 | 0.575 |  |
|  | MDA | 0.28 | 0.21 | 0.35 |  |
|  | HDA | 0.14 | 0.105 | 0.175 |  |
| b/tsDMARD 1 (late)<sup>3</sup> | Remission | 0.11 | 0.0825 | 0.1375 | (36) |
|  | LDA | 0.41 | 0.3075 | 0.5125 |  |
|  | MDA | 0.32 | 0.24 | 0.4 |  |
|  | HDA | 0.16 | 0.12 | 0.20 |  |
| b/tsDMARD2 (early)<sup>2</sup> | Remission | 0.12 | 0.09 | 0.15 | (37) |
|  | LDA | 0.30 | 0.225 | 0.375 |  |
|  | MDA | 0.39 | 0.2925 | 0.4875 |  |
|  | HDA | 0.19 | 0.1425 | 1.125 |  |
| b/tsDMARD 2 (late)<sup>3</sup> | Remission | 0.12 | 0.09 | 0.15 | (37) |
|  | LDA | 0.25 | 0.1875 | 0.3125 |  |
|  | MDA | 0.42 | 0.315 | 0.525 |  |
|  | HDA | 0.21 | 0.1575 | 0.2625 |  |

<sup>1</sup> Transition probability from one health state to another. All patients start at moderate disease activity.
<sup>2</sup> Early - drug is started after methotrexate failure
<sup>3</sup> Late - treatment after both methotrexate and triple therapy failure.

Laboratory costs were obtained from the hospital database. Costs of tests that were not offered at the hospital were obtained from private laboratories. We used three cost categories: costs at diagnosis, costs to start b/tsDMARDs, and costs to continue therapy (Table 2). Drug costs were obtained from the Tanzania Medical Stores Department (MSD) pricelist (29) which is the main drug supplier for the Ministry of Health. Where unavailable, we acquired wholesale prices from retail pharmacies. For the b/tsDMARD we opted for the cheapest options available in the market by contacting the companies providing them (Table 2).

Capital costs were obtained from the Mnazi Mmoja Hospital Engineering and Nursing departments (S2 Text). Costs of buildings, supplies and electricity were calculated per patient and combined as overhead costs (Table 2). We further assumed that the clinician, nurse, and pharmacist would each require 20, 10 and 5 minutes per outpatient visit, respectively. The personnel costs were obtained from the FairChoices DCP-4 analytic tool evidence brief for low-income country rates (30). All costs were point estimates. We used a range of ±50% for the univariate sensitivity analysis, and gamma distributions with 95% confidence intervals within these ranges for probabilistic sensitivity analyses (PSA).

### Effectiveness

Data on intervention effectiveness was obtained from previously published studies (31-37). When multiple studies on patients with RA for at least one year (established RA) reported CDAI outcomes, the data were pooled. The effects were translated into transition probabilities for each strategy, capturing the relative proportions of patients in the different health states at six months from treatment initiation (Table 1). Studies with established RA cohorts were used to reflect the Zanzibar Chronic Inflammatory Joint Disease (Zan-CIJD) cohort, where patients tend to have suffered relatively long before seeking care. More than 80% of patients who achieved the treatment target within the first six months remained in the target state receiving the same therapy over the next six months (37). For those on rescue therapy about 43% could reach treatment target at the end of each cycle.

Given the limited evidence, we made assumptions to determine some efficacy values. Once patients had exhausted all available treatment options within a strategy, they were categorized as either having reached the treatment target, or not, for the remaining cycles. Based on expert opinion, in the target group, patients were distributed in a ratio of 1:2 between remission and low disease activity, while in the non-target group, patients were distributed in a ratio of 2:1 between moderate and high disease activity.

We obtained mortality data specific to Tanzanian women from the World Health Organization’s health repository (38) and converted it into six-monthly mortality rates. To account for the higher mortality rate among patients with RA, we applied adjustments of 1.29 and 1.42 for low and moderate disease activity, respectively (39).

### Zanzibar study population

The Zan-CIJD study contains data collected from 1^st^ September 2019 to 28^th^ February 2022 from 102 patients with RA attending the rheumatology outpatient clinic at Mnazi Mmoja referral hospital. The majority were female (84%), with a mean age of 45 years (SD 13.5) and the mean disease duration was 6.4 years (CI 1.3). Their baseline CDAI was 19.8 (SD 12.8), indicating moderate disease activity.

### Utilities

Quality-adjusted life-years (QALYs) are a combined measure of the mortality and morbidity caused by a disease. They are calculated as a function of the time spent in a health state and the health-related quality of life (HRQoL) weight (i.e., utility score) associated with that health state (40). HRQoL utility scores for each level of disease activity were derived from the Zan-CIJD cohort using the EuroQol® 5 dimensions and 5 levels (EQ-5D5L) questionnaire. Data from 538 questionnaires were tabulated and converted to utilities using the healthy Ugandan population utility scores (41) for reference. At baseline, patients had a mean utility score of 0.62 (95% CI: 0.54,0.70).

In RA, HRQoL is mainly affected by disease activity regardless of therapy used (42, 43). Based on the CDAI scores for the Zan-CIJD population, patients were grouped into remission, low, moderate, and high disease activity. The average utility score for each of the groups was determined based on each health state (see Table 3).

**Table 3:** Utility values based on RA disease activity in the Zan-CIJD population.

| Disease activity (CDAI) | Health-related quality of life (utility score) | Range |  |
| --- | --- | --- | --- |
| Remission | 0.87 | 0.6525 | 1.0875 |
| LDA | 0.72 | 0.54 | 0.90 |
| MDA | 0.57 | 0.4275 | 0.7125 |
| HDA | 0.21 | 0.1575 | 0.2625 |
| Treatment target | 0.77<sup>1</sup> | 0.5775 | 0.9625 |
| Not at treatment target | 0.45<sup>2</sup> | 0.3375 | 0.5625 |

CDAI- Clinical Disease Activity Index, HRQoL- Health related Quality of Life, LDA – Low disease activity, MDA- Moderate disease activity, HDA- High disease activity

### Analysis

We report Incremental Cost-Effectiveness Ratios (ICERS), the ratio of the incremental costs divided by the incremental utilities gained (44). The baseline comparator used was methotrexate monotherapy. Discounting for costs and utilities was estimated at 3% per annum and half-cycle correction was done. We determined the willingness to pay (WTP) threshold at USD 228/QALY based on a recent study determining cost-effectiveness thresholds by Pichon-Riviere et al (45).

Univariate deterministic sensitivity analysis was performed for the key input parameters such as costs of drugs and probabilities of reaching treatment target. A range of ±25% of the point estimates was used to describe uncertainties around treatment effectiveness and associated utilities. We assumed wider ranges of ±50% to capture uncertainties around costs variables, which tend to show greater variation. We report the univariate analyses as a Tornado diagram.

We further performed probabilistic sensitivity analysis (PSA) using Monte Carlo simulation with 10000 iterations. We adopted the ranges from the univariate deterministic sensitivity analysis to conduct the PSA and assumed specific distribution shapes for parameters on costs (gamma), probabilities (beta) and utilities (normal). Incremental cost-effectiveness scatter plots for pairwise comparison of strategy 1 as base case analysis with strategy 2 and strategy 3 were also plotted. For visualization purposes we reduced the corresponding scatterplots to 750 iterations.

We performed the analysis with varying WTP-thresholds equal to one GDP per capita and twice the GDP per capita at USD 1136 and 2272, respectively. An analysis of the optimal treatment strategy using a WTP of USD 282 was also performed using Monte Carlo simulation. We also conducted price threshold analysis comparing methotrexate with the other treatment options to assess whether changes in drug prices would result in change in what is determined as optimal treatment strategy.

Finally, scenario analyses were done using drug costs from Scandinavian countries, and India (Table 4). Scandinavian drug costs were based on official prices for the csDMARDs and approximate costs for the two cheapest b/tsDMARDs in Norway for 2022 (drug names not given due to national tender requirements). Prices from India were obtained from the integrated pharmaceutical database site (46). If prices were unavailable, we selected the lowest retail prices from providers (47, 48).

**Table 4:**
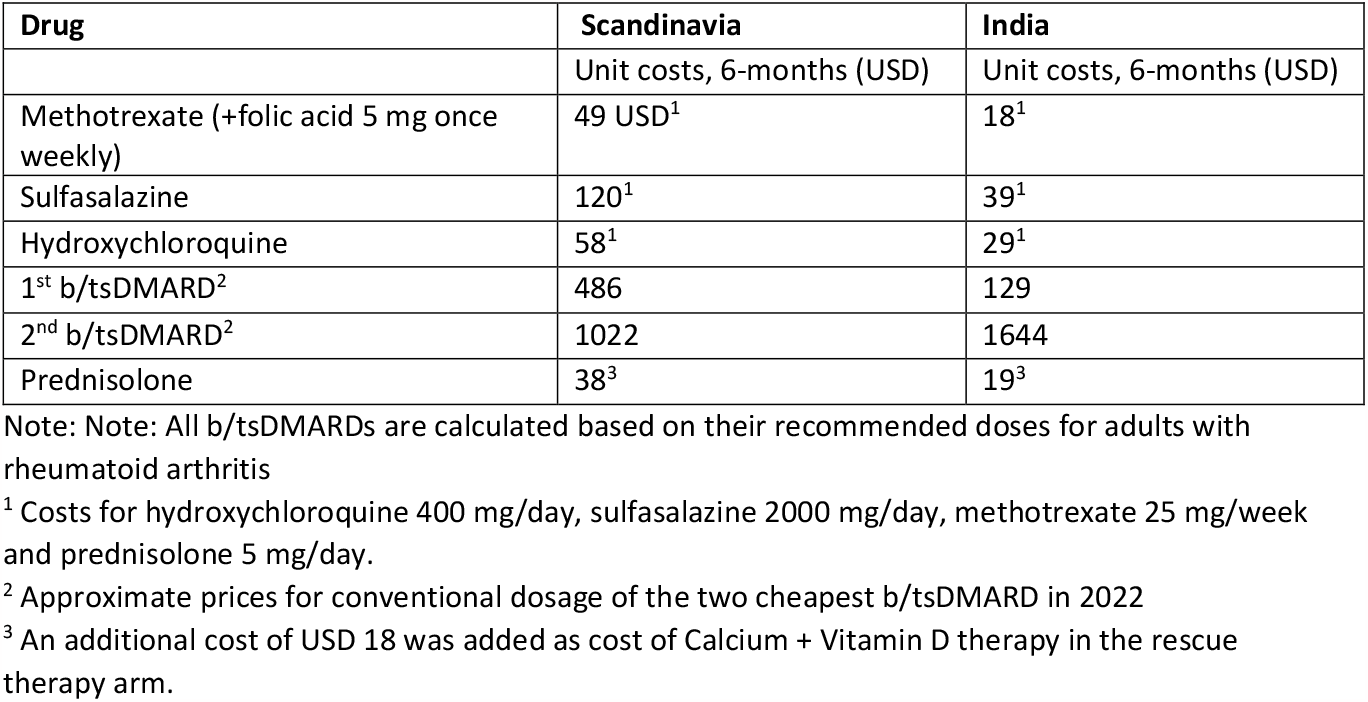
Drug costs for the management of RA from Scandinavia and India (six-monthly)

### Ethical considerations

Ethical approval was obtained from the Zanzibar Health Research Institute (ZAHREC/02/JULY/2019/43) and Norwegian Regional Committee for Medical Research Ethics (2019/472/REK vest) for the Zanzibar CIJD study. It included the use of data for this analysis. We obtained informed and written consent; illiterate patients provided their consent with a thumbprint.

## RESULTS

The difference in costs between the strategies were largely driven by the drug costs, with b/tsDMARDs costing significantly more than the conventional synthetic DMARDs. The lowest treatment cost was for strategy 1 (methotrexate therapy) at USD 634/patient/3 years and strategy six had the highest cost (methotrexate with two consecutive b/tsDMARDs) at USD 5,011/patient/3 years (table 5).

**Table 5:**
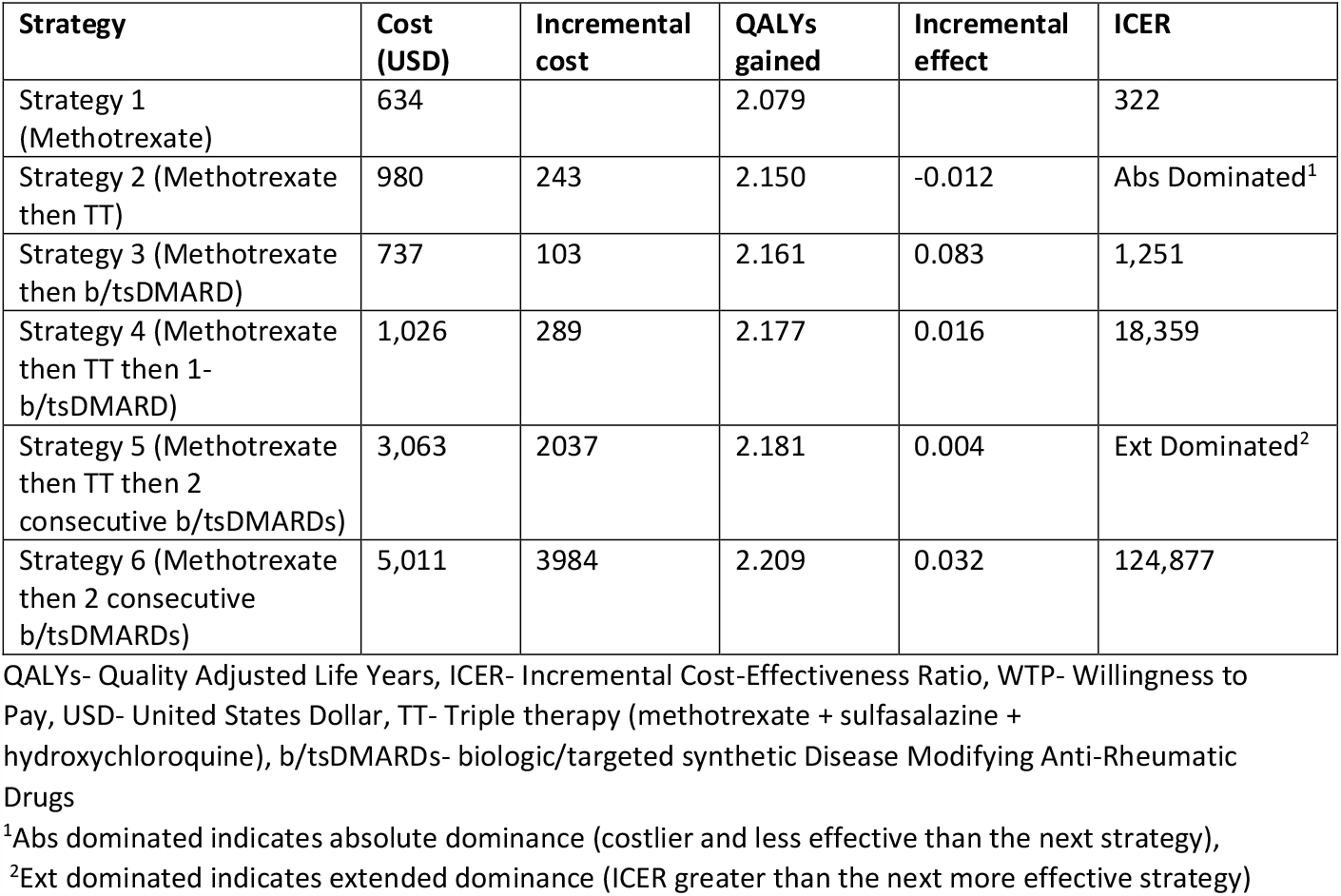
Three-year costs (mean USD per patient), effects (mean QALY gained per patient) and cost effectiveness of six treatment strategies for rheumatoid arthritis using Monte Carlo simulation with 10,000 iterations and WTP threshold of USD 282.

| Strategy | Cost (USD) | Incremental cost | QALYs gained | Incremental effect | ICER |
| --- | --- | --- | --- | --- | --- |
| Strategy 1 (Methotrexate) | 634 |  | 2.079 |  | 322 |
| Strategy 2 (Methotrexate then TT) | 980 | 243 | 2.150 | -0.012 | Abs Dominated<sup>1</sup> |
| Strategy 3 (Methotrexate then b/tsDMARD) | 737 | 103 | 2.161 | 0.083 | 1,251 |
| Strategy 4 (Methotrexate then TT then 1-b/tsDMARD) | 1,026 | 289 | 2.177 | 0.016 | 18,359 |
| Strategy 5 (Methotrexate then TT then 2 consecutive b/tsDMARDs) | 3,063 | 2037 | 2.181 | 0.004 | Ext Dominated<sup>2</sup> |
| Strategy 6 (Methotrexate then 2 consecutive b/tsDMARDs) | 5,011 | 3984 | 2.209 | 0.032 | 124,877 |

QALYs- Quality Adjusted Life Years, ICER- Incremental Cost-Effectiveness Ratio, WTP- Willingness to Pay, USD- United States Dollar, TT- Triple therapy (methotrexate + sulfasalazine + hydroxychloroquine), b/tsDMARDs- biologic/targeted synthetic Disease Modifying Anti-Rheumatic Drugs
<sup>1</sup>Abs dominated indicates absolute dominance (costlier and less effective than the next strategy),
<sup>2</sup>Ext dominated indicates extended dominance (ICER greater than the next more effective strategy)

The highest effectiveness amounted to 2.209 QALYs gained from treatment according to strategy 6 (methotrexate therapy + two consecutive b/tsDMARDs) while the lowest treatment effectiveness was obtained using strategy 1 (methotrexate alone), 2.079 QALYs (table 5).

From a Zanzibar healthcare provider perspective, for a WTP threshold of USD 282, strategy 1 was cost-effective while strategies 3, 4 and 6 were suboptimal options. ICERs ranged from 322 to 124,877 for all six strategies (table 5). Strategy 2 was found to be both more costly and less effective than strategy 3 and therefore not a rational choice from the healthcare provider’s perspective (absolutely dominated). Strategy 5 had an ICER greater than strategy 6 despite strategy 6 being more effective and was therefore excluded (table 5).

### Deterministic sensitivity analysis

One-way sensitivity analysis comparing the three strategies considerable under the set WTP thresholds (methotrexate monotherapy (strategy 1) compared to methotrexate + TT (strategy 2) and methotrexate+ b/tsDMARD1 (strategy 3)) showed that the most influential parameter on the ICERs was the cost of the TT and b/tsDMARD1 (Fig 3a and b). Compared to strategy one, the mean ICERs when considering single variables’ ranges remain largely above the WTP thresholds. However, comparing strategies 1 and 3, the assumed cost of the b/tsDMARD, at the lower end of its range, includes the option that strategy 3 is optimal in Zanzibar at the baseline WTP threshold, assuming all other variables constant (Fig 3b). Comparisons with other strategies found no single parameter had a significant effect on the ICER to cross the WTP threshold (S2-4 Fig).

**Fig 3.**
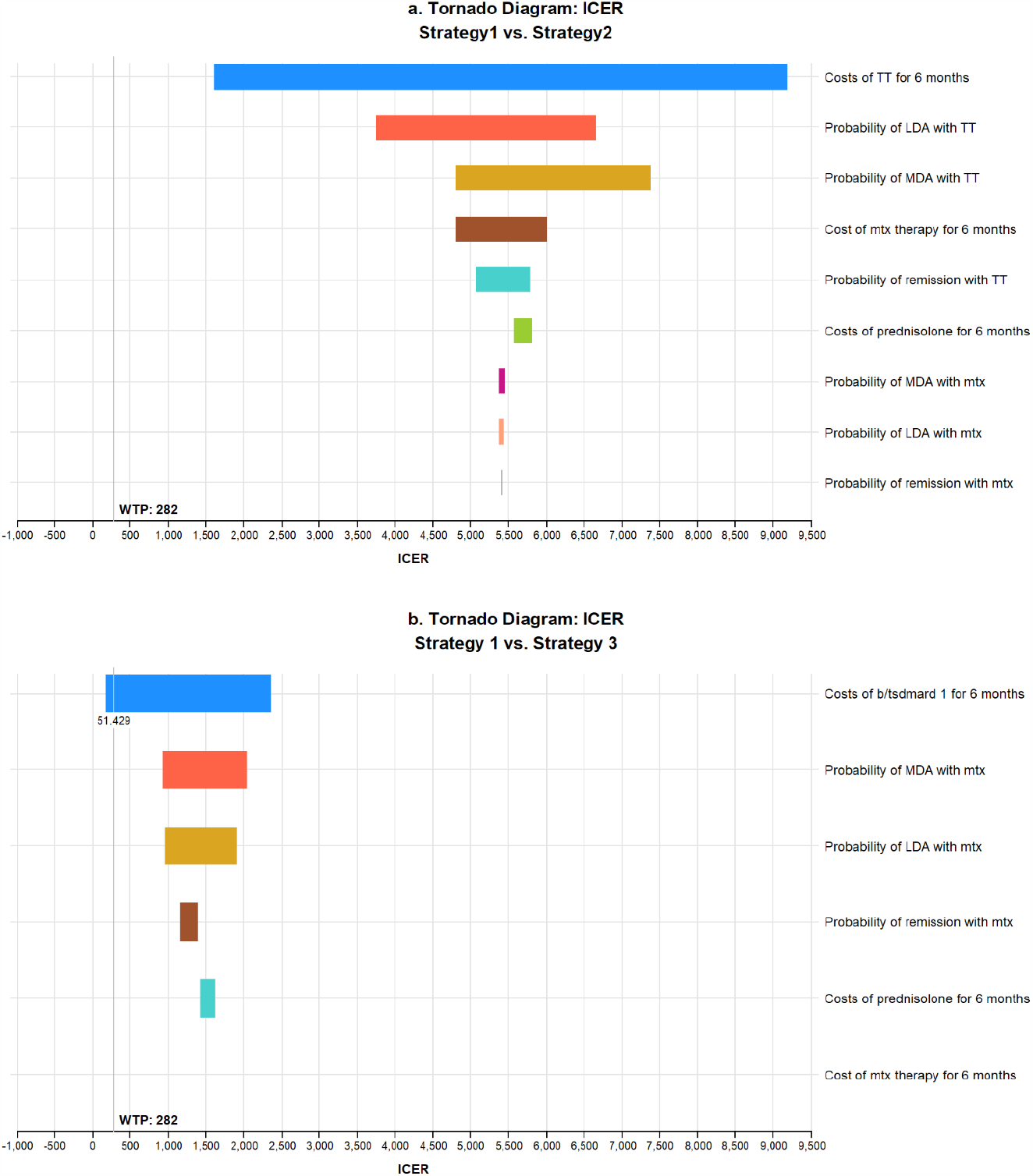
One-way sensitivity analysis comparing ICERS of methotrexate monotherapy vs methotrexate + TT (a) and methotrexate + b/tsDMARD1 (b). Each variable is arranged by decreasing impact on the ICER. A willingness to pay threshold of USD 282 was used.

### Probabilistic Sensitivity Analysis

Pairwise comparison of strategy 1 (methotrexate monotherapy) as the base case, with strategy 2 (methotrexate + TT) and strategy 3 (methotrexate+ b/tsDMARD1) showed that the iterations largely favored strategy 1 as the cost-effective option despite being less effective (Fig 4a and b).

**Fig 4a and b.**
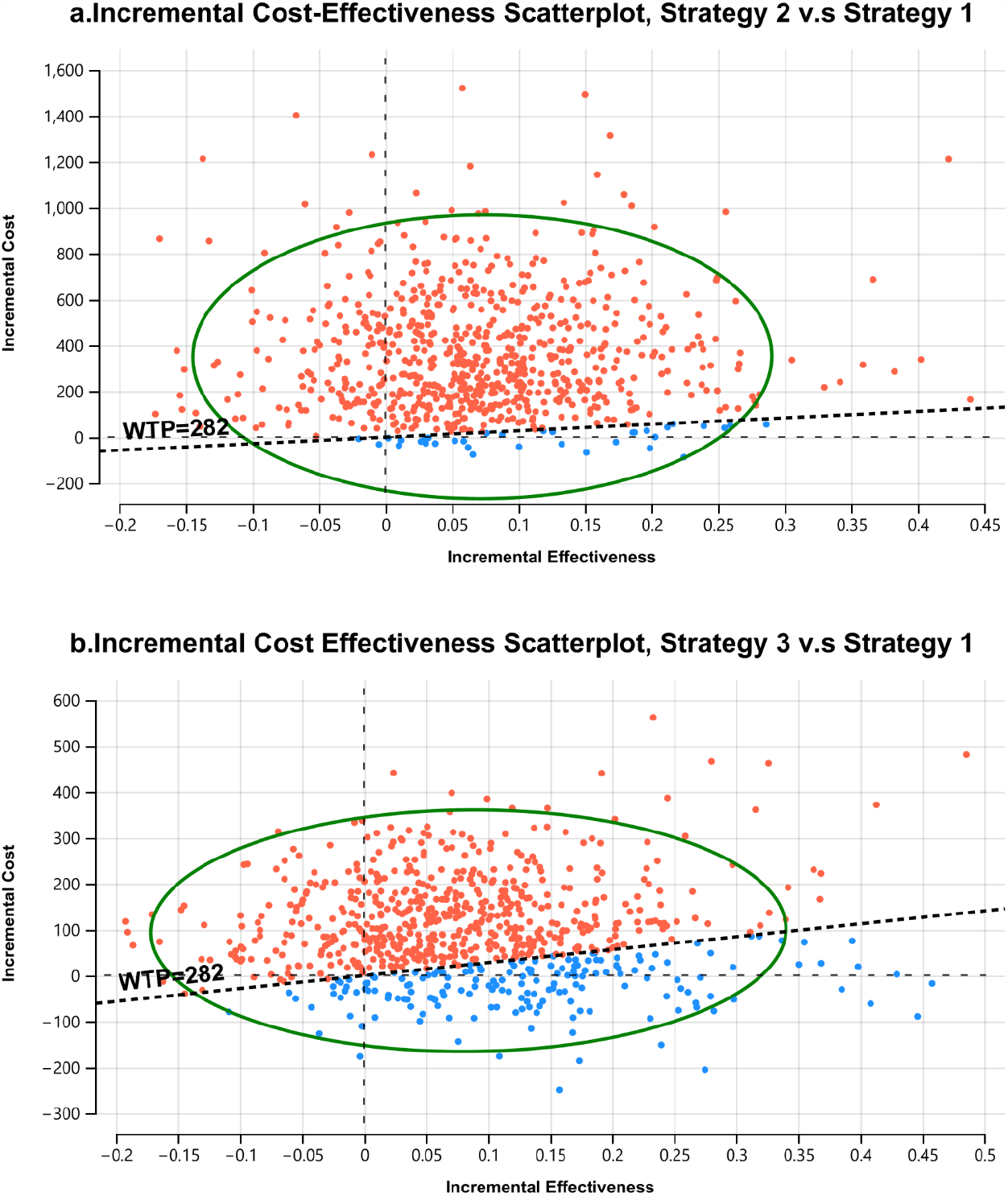
Incremental cost-effectiveness scatterplots, showing pairwise comparison of strategy 2 (methotrexate + TT, Fig 4a) and strategy 3 (methotrexate + b/tsDMARD1, Fig 4b), both compared to strategy 1 (methotrexate monotherapy) as the base case. The expected values for cost and effectiveness for strategy 1 are located at the intercept of the y and x axis, 0 cost, and 0 effectiveness. All iterations of strategies 2 and 3, shown as dots, indicate the incremental difference in cost and effectiveness to the average values for strategy 1. In most iterations, both strategies are more effective and more costly than the base case, indicated by the majority of iterations location in the North-East quadrant of the graphs. The dotted line delineates the willingness to pay threshold for Zanzibar (WTP) set at USD 282. With the estimated WTP, the more effective treatments are, with high probability, not cost-effective treatment options for Zanzibar, shown as red dots. While more costly and more effective, the incremental gain in effectiveness can hardly be justified with the corresponding incremental cost, when assuming the ICER threshold for Zanzibar. Only 4.7% of iterations suggested strategy 2 as cost-effective treatment in this comparison. Comparing strategy 3 to strategy 2, 23.5%, a higher share of iterations favor strategy 3. This suggests less certainty for a potential decision in favor of strategy 1 in this comparison with the uncertainty we introduced in the model.

The evaluation of the cost effectiveness when applying other WTP thresholds for Zanzibar of USD 1136 and 2272 indicated that with increasing WTP thresholds strategy 3 (methotrexate + b/tsDMARD) became a probable cost-effective treatment alongside strategy 1, and, less so, strategy 4 at the highest thresholds. (Fig 5).

**Fig 5.**
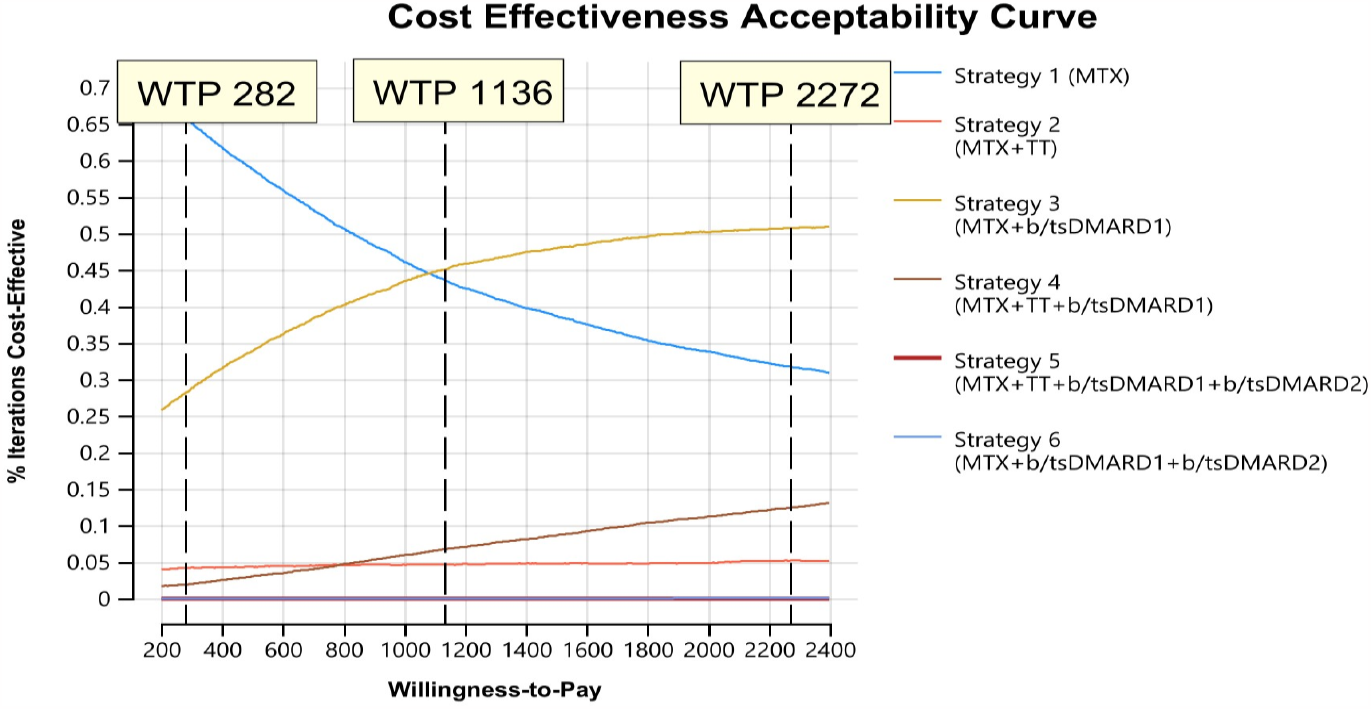
Cost effectiveness acceptability curve assessing feasible treatment strategies with variation in willingness to pay (WTP) thresholds.

Assuming the baseline WTP-threshold of USD 282, strategy 1 (72%) was likely the optimal strategy, with more than 20% of iterations favoring strategy 3 (Fig 5). Strategy 5 and 6 were not considered acceptable at any considered WTP.

At a WTP threshold of USD 282, price threshold analysis found that optimal prices for TT and b/tsDMARD1 were USD 102 and 61.5 respectively for the other treatment strategies to be considered cost-effective. There was no optimal price threshold for b/tsDMARD2 at a WTP of USD 282 (S1 Table).

### Scenario analysis

We performed two scenario analyses by including drug prices from Scandinavia and India in the Zanzibar-based model. Strategy 1 was still the probably cost-effective option in both, but due to global differences in prices, particularly for TT and the b/tsDMARDs, strategies 2,4 and 5 became considerable, though suboptimal, options in Zanzibar once price changes were assumed. Strategies 3 and 6 were always dominated (S2 Table).

## DISCUSSION

To our knowledge, this is the first study in Sub-Saharan Africa looking at the cost-effectiveness of multiple DMARD therapies. The study looked at the cost-effectiveness of providing treatment in a stepwise approach, which simulates a clinical setting and is in accordance with recommendations in international clinical guidelines. We do believe that using a T2T approach which predefines treatment goals and applies tight control via regular, appropriate treatment adjustment (49) enables limitation of costs as it is more effective in reaching treatment target than usual care (50). All patients are started on the most cost-effective drug and the least cost-effective therapies are reserved for few and select patients. The model is likely to be relevant in many African settings due to similarities across the region in delayed presentation and high disease activity among patients with RA (11,51,52).

In this study we found that methotrexate was the cost-effective RA treatment option in a Zanzibar setting. A systematic review and meta-analysis looking at cost-effectiveness of b/DMARD with csDMARDs as comparator also found that for LMIC, bDMARDs were not cost-effective (53). In high-income settings, bDMARDS were found to be cost-effective (6, 54), because of the higher cost-effectiveness threshold, particularly for those with low costs (55). Our findings suggest that when WTP is increased to USD 2272 equivalent to twice the GDP per capita then regimens with b/tsDMARDs become the optimal treatment strategy.

Most reference studies used to determine efficacies were from Western populations. In the Zanzibar setting, most patients are presented with established disease. For this reason, we included studies with patients with established RA and long-disease activity in our analyses, although there were relatively few studies for consideration.

LMICs tend to have higher drug prices compared to high-income countries (56). Currently, the cost of b/tsDMARDs makes them unfeasible for clinical use in a Zanzibar setting. However, the availability of biosimilar b/tsDMARDs for several TNF-inhibitors, as well as rituximab, has led to price reductions where they are accessible. Over time, this may increase the utilization of these drugs in LMICs. In Scandinavian countries, where the efficacies of b/tsDMARDs are considered comparable, costs play a significant role in the choice of which b/tsDMARD to use in therapy (29). As a comparison, the cost of triple therapy and bDMARDs was much lower in Scandinavian countries as compared to Zanzibar while csDMARDs were cheaper in India. To reduce costs in SSA, adopting confidential drug tendering schemes, as is common in Scandinavia (29) and other European countries (57), could be beneficial.

Institutionalisation of drug price negotiations is important to enable governments to negotiate discounted prices from the pharmaceutical companies, ensuring both cost-effectiveness and fair distribution globally (58). Additionally, neighboring African countries could explore bulk purchases to qualify for large discounts. In India, b/tsDMARDs are not included in the government standard treatment guidelines and patients purchase them privately which allows the pharmaceutical companies to have high prices.

With RA, early diagnosis and early initiation of treatment increases likelihood of response to therapy and slows disease progression. The majority of our patients had severe RA at presentation, with moderate to high disease activity when seeking hospital care. Programs to increase RA community awareness as well as health care provider training on early detection and T2T therapies could potentially improve patient outcomes (59). Additionally, this would help decrease the costs of care as well as prevent work disability which can be as high as 70%, 5–10 years after symptom onset (60). Furthermore, treatment costs for RA are at their highest at the time of diagnosis and it decrease over time (61).

Efficacy data used in the analysis were acquired from RCTs carried out in Western populations (30-35). They vary from our group in age of onset, disease duration and disease activity at presentation. There may also be discrepancies in therapeutic DMARD doses and treatment response in African populations compared to Western ones. Such factors may have influenced our findings and underline the need for more research on rheumatological conditions in SSA, particularly on the efficacy of DMARDs, to obtain reliable data from LLMIC settings. Such studies may also highlight the risk of b/tsDMARD side effects in resource-constrained regions where infectious illnesses, notably tuberculosis, are prevalent and pose a considerable risk to patients.

Furthermore, in most of the RCTs on RA, the ACR-20 is used as a measure of efficacy of therapy. However, ACR-20 is not a commonly used target in clinical settings. For this reason, we chose to use CDAI as disease activity measure, even though for majority of the trials, CDAI was only reported as a secondary outcome. This reflects a knowledge deficit regarding efficacy of treatment in clinical settings.

Despite the high costs of bDMARDs, rituximab (RTX) is available in Zanzibar for the treatment of cancer. The availability of RTX was largely due to political goodwill and interest in establishing oncology care on the island. With the government policy of free health care for all, this means that it is available to patients with rheumatic diseases as well, without prior assessment of rationale or fairness. Unfortunately, due to its high cost it is not cost-effective. With several effective b/tsDMARDs becoming widely available globally, this emphasizes the need for decision-makers to evaluate the cost-effectiveness of therapeutic options before they are made available in clinical settings. This also underlines the need for a universal health care package, particularly in resource-constrained settings, that determine which options give the best value for money as has recently been rolled out in the Zanzibaressential health care package (62) which contains DMARD combination therapy for RA treatment.

Cost-effectiveness is one of the most important criteria in health care priority setting. Our analysis indicates that methotrexate is the only feasible treatment option for patients with RA. However, our study showed that only around 15% would achieve the treatment target and remain on monotherapy. Given that RA is a severe disease, affecting HRqoL with pain over a long period of time, perhaps allowing for higher WTP thresholds are acceptable and strategy 3 is a more fair priority. This shows the limitations of a pure health maximizing perspective, and a Distributive Cost-Effectiveness (DCEA) study is perhaps more suitable. Although not explored by our analysis, other studies suggest that the high costs of care can be offset by the costs associated with lost productivity (63), which have a significant impact in RA patients because majority of patients are of working age. It is noteworthy that in SSA, for breast cancer therapy, costs of up to USD 20,000/QALY are considered acceptable in SSA (64), which is higher than the costs found for strategy 1, 3 and 4 in our study for RA treatment. Other options to consider for patients with RA requiring expensive drugs include tapering off expensive drugs and prioritizing only those with the most severe disease (65).

## CONCLUSION

Although b/tsDMARD therapy is considered optimal treatment in developed countries it is still not a cost-effective option in resource limited settings, mainly due to the high cost of drugs. Allowing for a higher WTP threshold would ensure that these options become more acceptable. Until they are made affordable, methotrexate monotherapy is considered cost-effective for the management of patients with RA in Zanzibar.

## Data Availability

The model and additional generated results are contained in the supplementary material.

## ACKNOWLEDGEMENTS

We acknowledge the contributions of many colleagues who helped in one way or another in finalizing this manuscript, in particular the registrars at the rheumatology clinic (Noorein A Omar, Faiza M Juma and Mariam A Mzee). Perez Ochanda provided valuable assistance with the utility scores and Jutta Jorgensen who gave valuable insight throughout the process.

